# Adolescent Normative Intervals for Body Surface Gastric Mapping: Spectral Analysis

**DOI:** 10.1101/2025.11.25.25340911

**Authors:** Gayl Humphrey, Gabriel Schamberg, Binghong Xu, Nicky Dachs, Daphne Foong, Chris Varghese, Christopher N. Andrews, Hayat Mousa, Armen Gharibans, Gregory O’Grady

## Abstract

**Objective:** Body surface gastric mapping (BSGM) non-invasively assesses gastric myoelectrical activity along with real-time symptom reporting. In adults, the development of normative intervals has underpinned new explanatory phenotypes, aiding clinical decision-making. This study established normative reference intervals for adolescent BSGM metrics.

**Study Design:** Healthy adolescents aged 12-17 years with a BMI <35 kg/m^2^ were recruited from New Zealand, Australia and the United States. BSGM using Gastric Alimetry (Alimetry, New Zealand) involved a 30-minute fast, followed by a 480-kcal meal, and a 4-hour postprandial recording. Reference intervals were calculated for four validated metrics: Principal Gastric Frequency (PGF), body mass index (BMI)-adjusted amplitude, Gastric Alimetry Rhythm Index (GA-RI, indicating rhythm stability), and the fed-to-fasted amplitude ratio (ff-AR). Results were reported at the median and 5th and/or 95th percentiles as appropriate.

**Results:** A total of 107 participants (52.8% female, median age 14 [IQR 13-16], median BMI 20.1 [IQR 18.75-22.40]) with mixed ethnicities were included. No substantive correlations were observed between BSGM metrics and demographics or anthropometric data. Therefore, a single set of normative reference intervals was established. Median PGF was 3.06 cycles per minute; reference interval 2.72-3.37. Median BMI-adjusted amplitude was 37.80 µV; reference interval 20.0-72.0. Median GA-RI was 0.51; reference interval ≥0.22. Median ff-AR was 2.12; reference interval ≥1.0.

**Conclusion:** This study presents normative reference intervals for BSGM spectral metrics in adolescent populations, informing the interpretation of tests in research and clinical practice.

## Introduction

Chronic upper gastrointestinal (GI) symptoms are common in children and adolescents.^1–4^ Despite their prevalence, the specific causes and underlying mechanisms of these disorders are poorly defined, which makes precise therapy challenging.^5,6^ Current diagnostic modalities in pediatrics include Rome Criteria^7^, antroduodenal manometry (ADM), and gastric emptying scintigraphy (GES). While these tests are widely deployed, many patients often do not receive a specific diagnosis based on underlying mechanisms, leading to trial-and-error care. New, less-invasive tests that can provide actionable biomarkers are urgently needed to advance care.

Body Surface Gastric Mapping (BSGM) using the Gastric Alimetry^®^ test (Alimetry, New Zealand) is a diagnostic test that measures the underlying myoelectrical activity which coordinates gastric motility^8,9^, together with validated, synchronised symptom and mental health profiling.^10–12^ BSGM has been shown to provide unique diagnostic insights beyond other investigations, such as ADM or GES, with superior symptom correlations, including in adolescent populations.^13–16^

A pilot study found that BSGM is feasible and safe in children as young as 5 years, but noted differences in BSGM metrics between children and adolescents.^17^ Similarly, while early studies suggested that core BSGM metrics in healthy adolescents were comparable to adult normative reference intervals, some differences were observed, particularly in the rhythm stability (GA-RI) metric. These observed variations are likely attributable to the unique developmental and physiological characteristics distinguishing children, adolescents, and adults.^18,19^ They also emphasise the need for age-specific normative reference ranges that can be used to support the clinical interpretation of BSGM tests, such as those already established in the adult population aged 18 years and older.^20,21^

Therefore, this study aimed to establish normative reference intervals for adolescents aged 12-17 years for key spectral metrics of the Gastric Alimetry System, specifically the Principal Gastric Frequency, Gastric Alimetry Rhythm Index (GA-RI), body mass index (BMI)-adjusted amplitude, and the fed-to-fasted amplitude ratio (ff-AR). These normative intervals will be applied to guide clinical interpretation and provide clarity regarding biological variability in adolescents’ gastric myoelectrical activity.

## Methods

This was an observational cohort study conducted in Auckland (New Zealand), Philadelphia (USA), and Western Sydney (Australia), and ethical approvals were obtained from the relevant IRBs (references 2022 Full12705, 2024-PR12709, REB19-1925 and 21-018520, H15157). All enrolled participants provided assent or consent, and, where applicable, parent consent and / or assent. The study is reported in accordance with the STROBE statement.^22^

### Population

Adolescent healthy control participants aged 12-17 were recruited through local advertising and networks between 2023 and 2025. Interested participants were screened and excluded if they had active or a significant history of GI symptoms, were on prescribed medications known to affect gastric motility, including opioid analgesics, antiemetics and anti-spasmodics, had previous upper GI surgery or were positive for a gastroduodenal disorder after completing the Rome IV questionnaire.^7^ Patients with BMI >35 kg/m^2^, active abdominal wounds or abrasions, fragile skin, and adhesive allergies were excluded.

On completion of the Gastric Alimetry test, additional exclusion criteria were applied to mitigate the confounding effects of subclinical gastroduodenal disorders on test results. Participants were excluded from the final analysis if they met either of the following conditions upon completion of the Gastric Alimetry test: (1) Reported average severity of any individual continuous symptom exceeded 3/10; or (2) Reported an excessively full score >1/10 at the three- or four-hour postprandial period. These conservative inclusion criteria were deemed appropriate in view of the known burden of subclinical GI disorders in the general population.^21^ A similar approach was applied in the published BSGM adult normative intervals.^20^ Participants were also excluded if greater than 50% of the recording was affected by artifacts.^23^

Enrolment demographics were continuously monitored during recruitment to ensure capture of diverse ethnicities and age ranges.

### Gastric Alimetry System

The Gastric Alimetry System comprises a high-resolution, stretchable array embedded with 64 electrodes (an 8x8 array with an inter-electrode spacing of 1cm) and a wearable reader. An App on iPadOS guides setup and anthropometric measures, enabling personalised array placement on the abdomen (Figure 1).^10^ Within-test symptom logging utilises co-designed and validated pediatric pictograms.^24^ More details on the Gastric Alimetry system can be found elsewhere (refer^8,9,23,25^).

**Figure 1.**
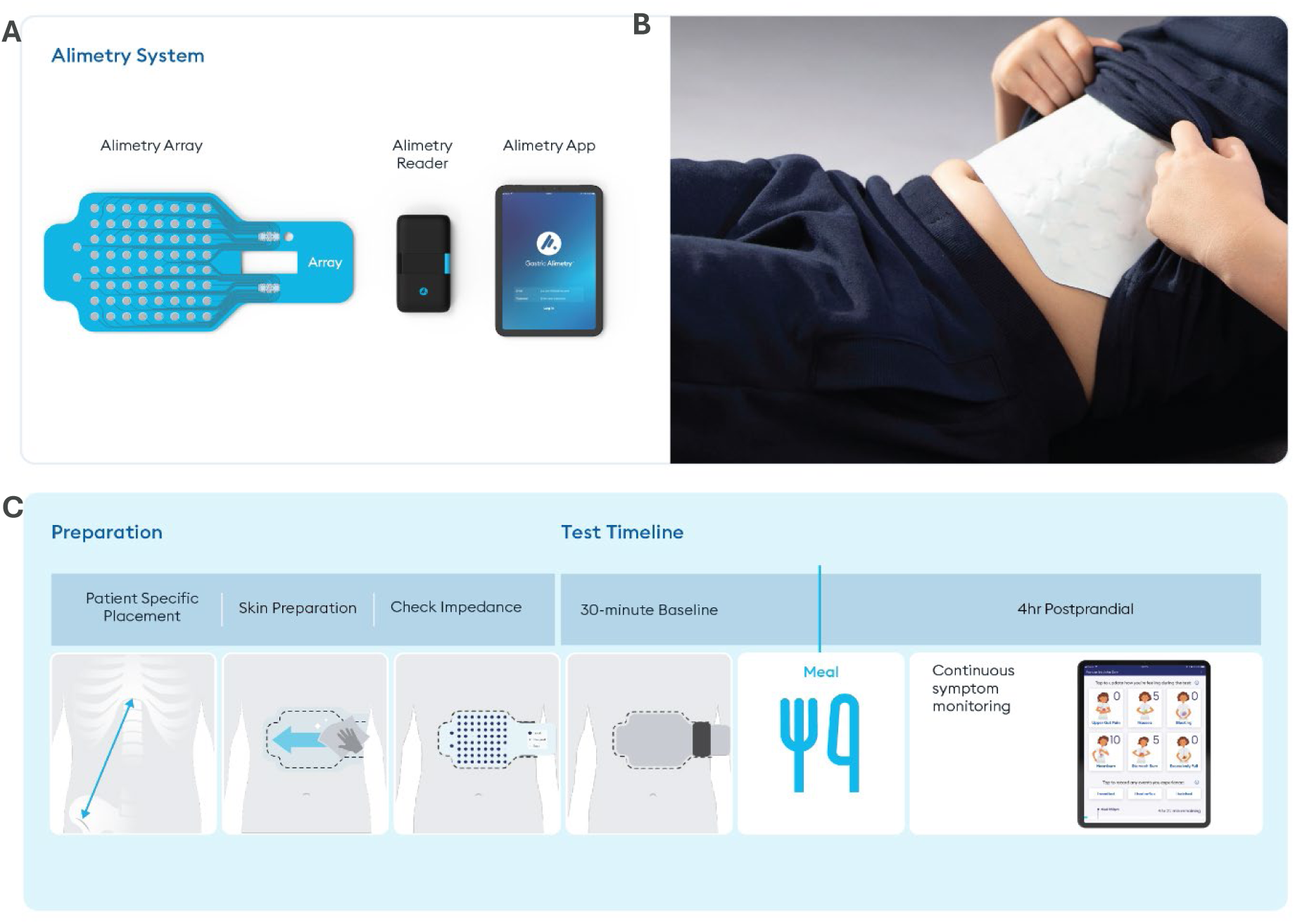
A. Gastric Alimetry System**, B.** The array placed on the abdomen of an adolescent, and **C.** the BSGM Standard Protocol illustrating the preparation phase and the elements in the test timeline [baseline, meal and continuous symptom reporting]

### Study Protocol

This study followed the standard Gastric Alimetry BSGM protocol (Figure 1).^9^ Participants fasted for at least six hours before the test. Personalised measurements and skin preparation were then completed before the array was placed. Next, participants were seated comfortably and reclined at a 45° angle. A 30-minute baseline recording was obtained, followed by a 480-kCal meal comprised of a nutrient drink (9g fat, 34g carbohydrate, 9g protein) and an oatmeal energy bar (5g fat, 45g carbohydrate, 10g protein, 7g fibre) consumed within 10 minutes, and immediately followed by a 4-hour postprandial recording. Participants were instructed to limit movement, talking, and sleeping, although they could mobilise for comfort breaks, and could read, watch TV, or work on a mobile device if desired. Although this protocol was designed for use with adults, it has been reported to be feasible, acceptable, and safe for children as young as 5 years old.^17^

### BSGM Data

Data were analysed using the Gastric Alimetry Algorithm v3.0.0, comprising a collection of signal processing steps including filtering, automated artifact correction, channel selection, and spectral analysis. The primary outputs are a spectrogram and the associated spectral metrics (PGF, GA-RI, BMI-Adjusted Amplitude, and ff-AR) reported for the entire test duration, as well as for the preprandial period and each postprandial hour. The development and validation of these metrics in adults have been well described elsewhere (refer ^20,21^); however, a brief outline of each metric is presented below, with a corresponding visualization presented in Supplementary Figure 1.

#### Principal Gastric Frequency (PGF)

PGF is reported in cycles per minute (cpm) and is the measurement of the sustained frequency within the ‘’plausible’’ gastric range, to avoid influence from motion artifacts and periodic adjacent colonic activity. PGF is not reported when there is no identifiable stable and coordinated gastric activity, as measured by GA-RI (see below).

#### BMI-Adjusted Amplitude

Measured in microvolts (µV), amplitude represents the total electrophysiological activity (both slow waves and smooth muscle contractions) detected at the body surface.^9^ It is calculated as the maximum amplitude (across frequency) for each time point and reported as a time-averaged value. Because average amplitude has a nonlinear correlation with BMI^8^, a BMI correction is applied using a multiplicative regression model to improve the utility of the amplitude reference interval.

#### Gastric Alimetry-Rhythm Index

This is a recent metric introduced as an alternative to previous instability coefficients reported in legacy electrogastrography studies^25^ that quantifies the concentration of activity within the principal gastric frequency band relative to the rest of the spectrum over time. A high GA-RI indicates a distinct, stable slow wave band, whereas a low GA-RI signifies a high degree of spectral scatter. GA-RI is correlated with BMI and is adjusted using a linear regression model and subsequently scaled to a value between 0 (lowest stability) and 1 (highest stability).

#### Fed-to-Fasted Amplitude Ratio

ff-AR is designed to capture the increase in gastric signal amplitude that typically occurs after a meal. Instead of using a single short postprandial window, the ff-AR is calculated as the ratio of the maximum 1-hour average postprandial amplitude to the pre-meal average amplitude. This ratio primarily reflects the initiation of contractile activity in response to eating. ff-AR is reported only as an overall value for each participant. The potential for a high fasting baseline has been shown in the adult data^23^; therefore, this metric is used only as a supporting indicator for an abnormal test in combination with other metrics.

### Statistical Analysis

Before analyses, skewed data were log-transformed. Univariable associations between participant characteristics (age, sex, BMI, and ethnicity) and spectral BSGM metrics (PGF, BMI-adjusted amplitude, GA-RI, and ff-AR) were assessed. Univariable regression models were used for continuous variables (age and BMI), a t-test for categorical variables (sex), and ethnicity was analysed using one-way ANOVA. Regression models were used for BSGM metrics, reporting standardised coefficients (mean-centred and standardised beta coefficients) due to the different unit scales of variables.

The estimated median and 5th and 95th percentiles for all metrics using the median-unbiased estimator for the whole cohort and stratified by sex and age group. 90% confidence intervals (CIs) were computed for each estimate using a nonparametric bootstrap method with 1000 bootstrap samples.

Cross-validation is a common technique used to evaluate a statistical model’s ability to generalise to new, unseen data. In the context of normative reference intervals, this means confirming that an independent sample of healthy controls will have the expected percentage of metrics falling outside the established range. A five-fold cross-validation analysis was used to test the applicability of each reference interval to participants not included in the interval calculation (external participants). The complete cohort was randomly partitioned into five folds, each consisting of 22 participants (Folds 1 and 2) or 21 participants (Folds 3-5). One group served as the test set, while the remaining four groups were used to compute the reference interval.

The normative reference intervals were then determined by rounding the 5th percentile for the one-sided reference intervals (ff-AR and GA-RI) and rounding the 5th and 95th percentiles for the two-sided intervals: BMI-adjusted amplitude and Principal Gastric Frequency.

Adolescent physiology is characterised by continuous growth and development, necessitating special consideration of the statistical approaches to accurately define "normal" ranges across healthy adolescents aged 12-17 years. Adolescent age-related developmental changes vary due to the complex interplay of genetic inheritance, environmental factors, such as nutrition and socio-economic status, and hormonal fluctuations unique to the pubertal process.^18,19,26^ As a proxy for evaluating potential age-related physiological differences, the cohort was also partitioned into two distinct subgroups: 12-14 years and 15-17 years and analysed. The significance level was set at α = 0.05 for all statistical analyses.

### Sample Size

A cohort of 107 healthy adolescents was used to establish normative reference intervals for BSGM spectral metrics. The sample size is consistent with several studies that have developed normative reference intervals in GI motility.^27,28^ Adequacy of the sample size was further assessed using bootstrap and cross-validation analyses, as described above.

## Results

A total of 123 healthy controls aged 12-17 years completed Gastric Alimetry (Refer to Supplementary Figure 2 for the study recruitment flow diagram). Of these, 16 (13%) participants were excluded for exceeding the symptom threshold during testing (score > 3; n=6 [4.8%]) and/or for excessive artifacts (>50%; n=10 [8.1%]). There were no demographic differences between included and excluded participants (all p > 0.05).

The demographics of the 107 resultant participants are reported in Table 1. Fifty-six (52.8%) were female, the median age was 14.0 years (IQR 13.0-16.0), the median BMI was 20.10 kg/m² (IQR 18.9-22.4), and participants encompassed a diverse range of ethnicities.

**Table 1.** Participant Characteristics

| Characteristics | Overall<br>(N = 107) |  |
| --- | --- | --- |
| **Sex: Female** [N (%)] | 58 (54.2) |  |
| **Age** [Median (min-max)] | 14.00<br>(12.00 - 17.00) |  |
| **Age Years** [N (%)] | **Female** | **Male** |
| 12 | 11 (52.4) | 10 (47.6) |
| 13 | 6 (42.9) | 8 (57.1) |
| 14 | 13 (56.5) | 10 (43.5) |
| 15 | 13 (68.4) | 6 (31.6) |
| 16 | 12 (63.2) | 7 (36.8) |
| 17 | 3 (27.3) | 8 (72.7) |
| **Height (cm)** [Median [IQR] | 166.0<br>(158.25 - 175.00) |  |
| **BMI kg/m<sup>2</sup>** [Median [IQR] | 20.10<br>(18.82 - 22.40) |  |
| **Ethnicity** [N (%)] |  |  |
| African / African American | 4 (3.7%) |  |
| American Indian/Alaskan | 1 (0.9%) |  |
| Asian | 9 (8.4%) |  |
| European / White | 74 (69.2%) |  |
| Indian / Sri-Lankan | 5 (4.7%) |  |
| Māori* and Pacific Islander | 10 (9.3%) |  |
| Middle Eastern | 2 (1.9%) |  |
| Other | 1 (0.9%) |  |
| Declined | 1 (0.9%) |  |

\*Māori: Indigenous peoples of New Zealand

Participants were distributed across the 12-17 age years and had an approximately balanced representation of sexes, as reported in Table 1.

As some metrics are BMI-adjusted, demographic characteristics (age, sex, ethnicity) were assessed for their relationship to anthropometric measures (height and BMI). There was a weak to moderate positive relationship between age and BMI (r = 0.25; CI 0.06-0.42, *p =* 0.009) only. Meal completion was consistently high (median 100%; IQR 80-100%), with no significant difference between females (median 90%; IQR 80-100%) and males (100%, IQR 75-100%); p = 0.37. Older adolescents (15-17 years) showed a significant trend toward slightly higher meal completion rates (r = 0.23; p = 0.017).

The median percentage of artifacts during testing was generally low (median 13.87%, IQR 8.8-30.5), with no differences observed by age (p = 0.31), age grouping (12-14 vs 15-17 years; p = 0.15) or sex (p = 0.21).

As expected for a control sample, the median total symptom burden score was very low at 1.5/70 (IQR 0.0-4.5), although this was slightly higher in females than males (2.1 (0.6-5.8) vs 0.4 (0.0-3.1); p=0.005). There were no significant differences in total symptom scores by age groups (p = 0.78) or BMI (p = 0.15).

### Associations between Demographics, Anthropometric Variables, Symptom Burden, and BSGM Metrics

Correlations were assessed between participant characteristics (demographics, anthropometrics, and total symptom burden scores) and the four BSGM metrics. The primary finding was that multivariate models, which tested the combined predictive power of all characteristics, did not significantly predict overall BSGM metric outcomes.

Several individual (univariate) negative correlations were observed between anthropometric variables and BSGM metrics. Age and height were both found to be negative predictors of Principal Gastric Frequency in the univariate analyses (r = -0.26, p = 0.007 and r = -0.29, p = 0.003, respectively); however, when included in the multivariate model, age was not a significant independent factor (r = -0.16, p = 0.15). BMI showed a negative association with two distinct BSGM metrics: BMI-adjusted Amplitude (r = -0.27, p = 0.005) and the GA-RI (r = -0.21, p = 0.03). This suggests that, although these metrics are both modestly adjusted for BMI, a weak trend towards decreasing values for BMI-adjusted amplitude and GA-RI remains with higher BMI. Finally, the fed-to-fasted amplitude ratio (ff-AR) was negatively predicted by age (r = - 0.20, p = 0.043), BMI (r = -0.27, p = 0.005), and height (r = -0.23, p = 0.016). In the multivariate model, only BMI remained a significant independent predictor of ff-AR (r = -0.22, p = 0.03). No individual associations were observed by ethnicity or symptom burden. These findings are presented visually in Figure 2 and detailed in Supplementary Table 1.

**Figure 2.**
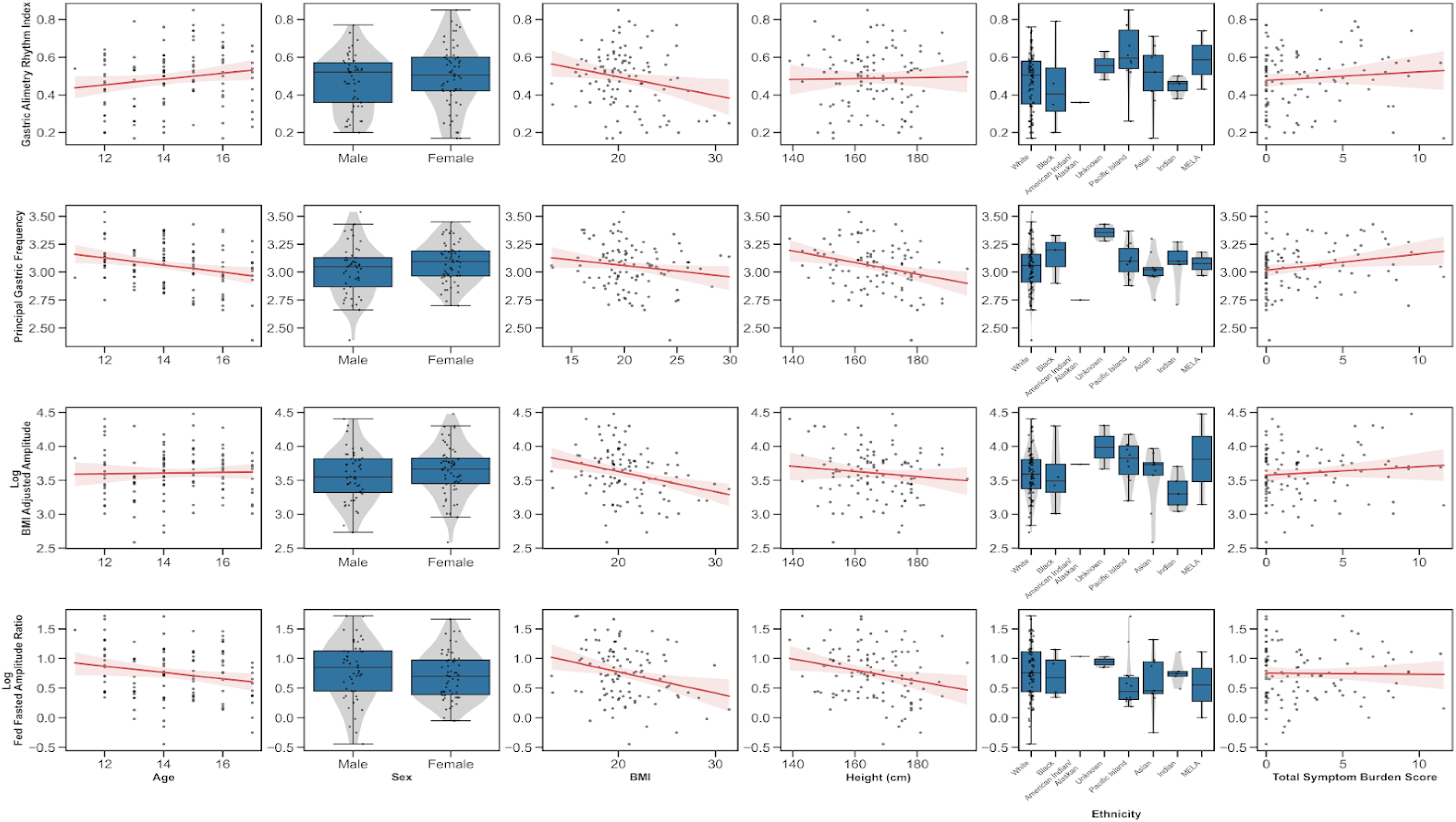
Association between patient demographic characteristics (age, sex, height, BMI, ethnicity) and total symptom burden and BSGM metrics (GA-RI, PGF and log-transformed BMI-adjusted amplitude and ff-AR).

The average spectrograms and BMI-adjusted amplitude curves for the total healthy control cohort, as well as by sex and age groups (12-14 and 15-17 years), are shown in Figure 3. All spectrograms visually demonstrate a stable PGF and a strong GA-RI, with a positive increase in amplitude immediately post-meal, and minimal qualitative differences, matching the quantitative analysis presented in Figure 2 and detailed in Supplementary Table 1. These features are characteristic of gastric myoelectrical function and define the normal BSGM spectral analysis in adolescents.

**Figure 3.**
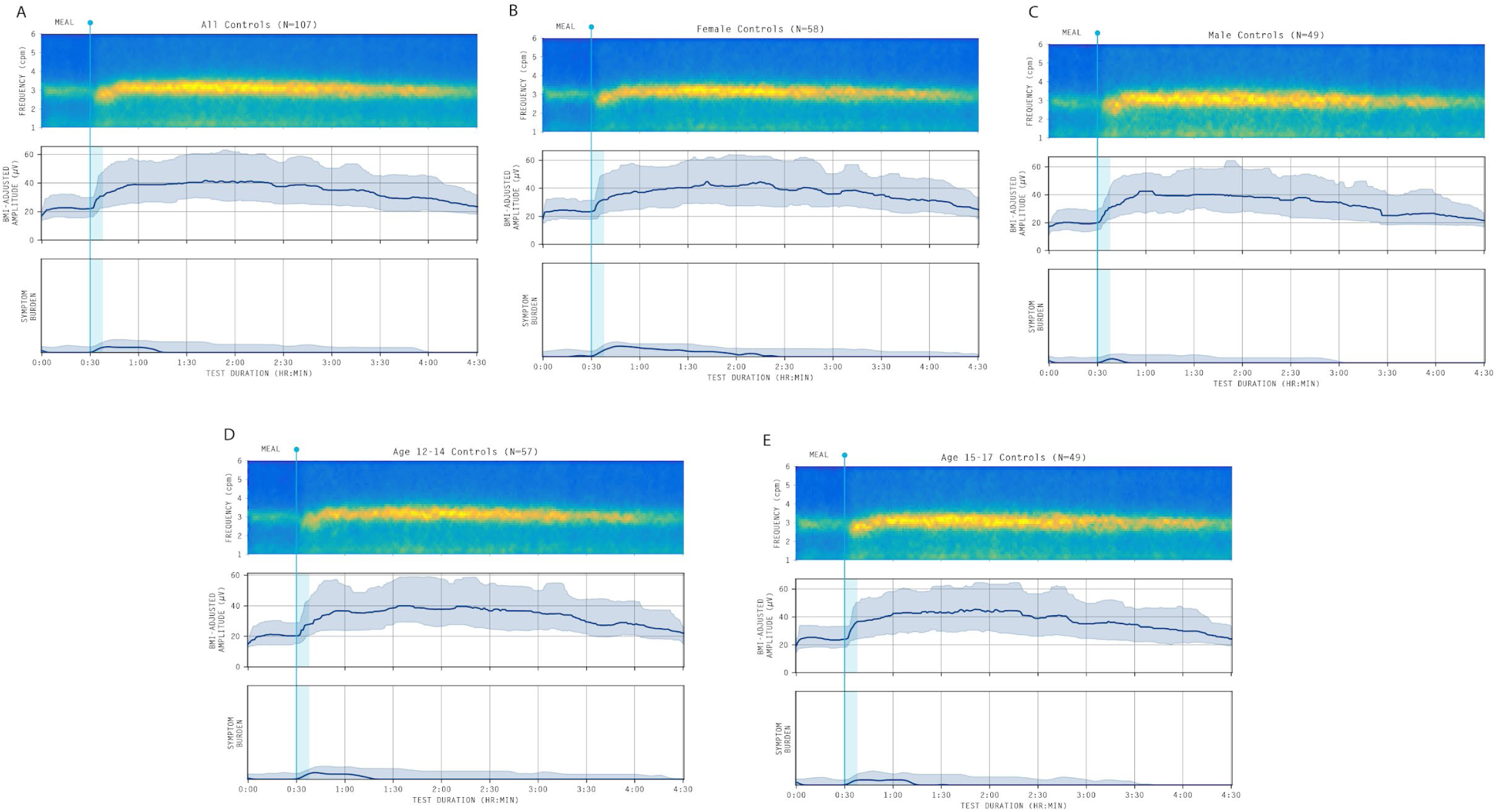
Averaged spectrogram and median BMI-adjusted amplitude curve with shaded interquartile range for (A) Whole cohort, (B) Female participants, (C) Male participants, (D) aged 12-14-year cohort and (E) aged 15–17-year cohort. All amplitude curves are displayed after applying a median filter for visual clarity.

No differences were observed in any of the average BSGM metrics between the sexes or by age (in years). PGF was higher in the younger-aged 12–14-year-old cohort (median 3.11 cpm, CI 2.96−3.21) compared to the cohort aged 15-17 years (median 3.00 cpm; CI 2.82−3.13; p=0.01). Despite reaching statistical significance, the magnitude of this difference (0.11 cpm) is unlikely to be clinically meaningful or alter a diagnostic conclusion.

BMI-adjusted amplitude exhibited a strong response to the meal challenge from median amplitude 22.4 μV pre-meal to median amplitude 39.22 μV post-meal (p < 0.001), with no differences observed based on age (in years), age groups, sex, BMI, or the percentage of the meal consumed (adjusted p>0.05 for all comparisons).

#### Normative reference intervals for BSGM metrics

The median, 5th percentile, 95th percentile and associated 90% bootstrap confidence intervals (CI) for each BSGM metric, computed overall and stratified by sex, are summarised in Table 2. The median and IQR for BMI-adjusted amplitude, PGF and GA-RI for specific BSGM test time intervals (pre-prandial and each one-hour window postprandially and by sex and age subgroups) are reported in Supplementary Table 2.

**Table 2.**
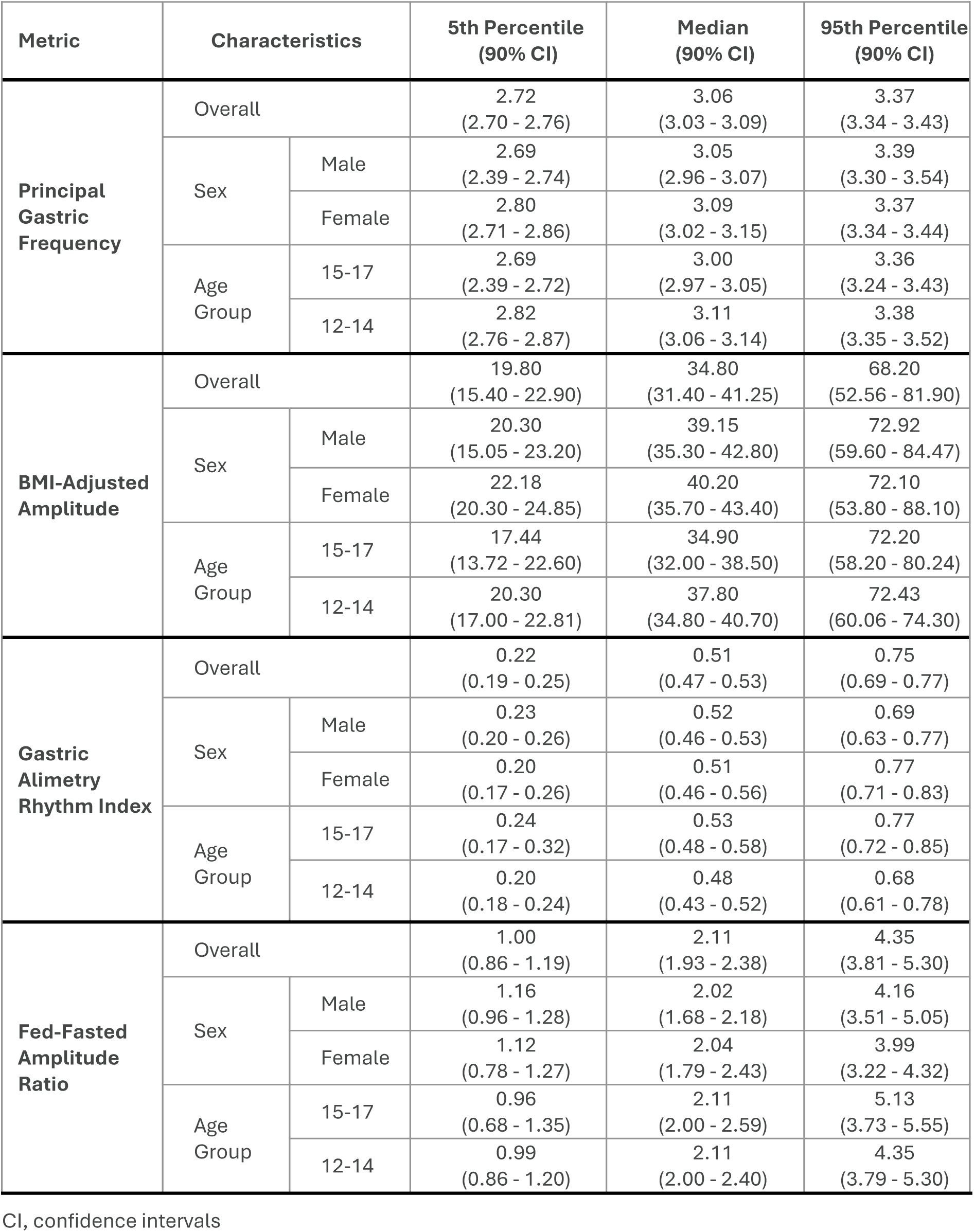
Normative adolescent healthy control data reporting median, 5th, and 95th percentiles for PGF, BMI-adjusted amplitude, GA-RI and ff-AR, with associated 90% CIs for the overall adolescent cohort and stratified by sex and age cohort for BSGM metrics

As the normal physiological gastric response was not meaningfully dependent on demographic or anthropometric factors (see above), a single, unstratified set of BSGM normative reference intervals was determined, as presented in Table 3.

**Table 3.** Adolescents aged 12-17 years Normative Reference Intervals for BSGM metrics

| BSGM metric | Lower | Upper |
| --- | --- | --- |
| Principal Gastric Frequency | 2.72 | 3.37 |
| BMI-Adjusted Amplitude | 20 | 72 |
| Gastric Alimetry Rhythm Index | 0.22 |  |
| Fed to Fasted Amplitude Ratio | 1.0 |  |

#### Cross-validation Analysis

Cross-validation analyses revealed that for each test group, we can expect 5% of independent healthy adolescent controls to fall outside the interval for ff-AR and GA-RI, and 10% to fall outside the two-sided intervals for BMI-adjusted amplitude and PGF (Supplementary Table 3). Supplementary Figure 1 presents a sample of individual spectrograms that fell outside the reference intervals for each metric.

#### Meal Response Ratio

While not currently a formal BSGM metric, the meal response ratio (MRR) is an exploratory measure being used to evaluate and visualise the temporal dynamics of the postprandial gastric response in clinical studies.^14^ It is expected that the ratio will be greater than 1.0 when comparing the first 2 hours post-meal to the subsequent two hours, reflecting the domain peak of myoelectrical activity during the initial phase of digestion. The median MRR was 1.21 (IQR 0.99-1.57) for the whole cohort. Groupwise examination showed that MRR was slightly higher in males (1.30 (1.05-1.52) compared to females (1.17 (0.97-1.51) with a similar result observed by age cohorts (12-14 years (1.28 (1.05-1.55) v 15-17 years 1.11 (0.97-1.62)), however, these differences were not statistically significant (p values 0.13 and 0.82 respectively).

## Discussion

This study characterised and defined normal gastric myoelectrical activity using BSGM in a healthy adolescent population aged 12-17 years and presents a set of normative reference intervals for Gastric Alimetry testing. As minimal demographic or anthropometric differences were found between age, age groups, sexes, and ethnicities, a single unified reference range could be formulated, which will now be fundamental in interpreting BSGM test data in clinical practice for adolescents.

Legacy electrogastrography studies highlighted the potential of gastric electrophysiology, including in pediatric populations, but methodological flaws and a lack of standardisation prevented their clinical adoption.^25,29,30^ BSGM represents a significant technological advance, providing a validated high-resolution protocol and superior signal processing pipeline that accurately reflects gastric activity.^21,23^ This is clinically important, as early BSGM studies have already demonstrated the ability to differentiate adolescents into distinct motor (e.g., dysrhythmic, weak meal responses) versus sensory phenotypes, where symptom-based diagnoses such as functional dyspepsia and gastroparesis overlap.^16^ Those studies identified ’abnormal’ activity; however, their findings cannot be confidently applied in clinical practice without a clear definition of ’normal’. The normative reference intervals established in this study provide an essential foundation that now enables clinicians to rigorously distinguish these pathological phenotypes from normal physiological variation in adolescents.

The emerging BSGM phenotypes are demonstrating clinical relevance. For example, the dysrhythmic phenotype, primarily defined by a low GA-RI but potentially accompanied by a low amplitude, has been observed in both adult and pediatric patients in association with nausea and vomiting syndromes, with improved symptom correlations versus delayed gastric emptying.^31–33^ This spectral pattern is considered indicative of a neuromuscular disorder (e.g., ICC network injury or dysfunction), and recent work has correlated low GA-RI values with neuropathic findings on ADM.^15,34^ The reference interval for GA-RI (≥0.22) established in this study now provides an objective, non-invasive threshold to help identify this clinically significant phenotype in adolescents, potentially sparing some from more invasive investigations.^16^

This study also provides data on the normal meal response, another valuable diagnostic feature. The cohort generally exhibited the expected positive amplitude increase post-prandially (median ff-AR 2.12). While some participants showed a high fasting baseline that blunted their individual ff-AR and the reference interval, the group-level data (Figure 3) confirms that a robust meal response is typical. However, as observed in the adult reference intervals, the high variability of this metric among healthy controls means that it is less clinically valuable in isolation, and generally serves as a supporting measure rather than having primary diagnostic importance.^23^ In addition, our exploratory MRR metric (median 1.21) offers a potential alternative method for quantifying meal responses in the future and may help predict delayed gastric emptying and/or impaired gastric accommodation as seen in adults^32^, although further research is required before this can be considered a clinical Gastric Alimetry test parameter.

Overall, the physiological consistency confirms that a single normative range can be applied to adolescents across the 12– to 17-year age span, regardless of their demographic or anthropometric differences. Additionally, the cross-validation method enables an estimate of the extent to which the percentage of healthy controls from an independent cohort is expected to fall outside the reference intervals. The findings support the validity and generalisability of these adolescent reference intervals for assessing abnormalities in clinically applied BSGM tests.

Some limitations to this study are acknowledged. While BSGM has been found to be safe and acceptable in children as young as 5 years^17^ (not studied here), the exclusion of adolescents with test artifacts >50% means caution is warranted when interpreting BSGM clinically with high artifacts. The 4.5-hour test protocol, although comparable to gastric emptying scintigraphy, is lengthy and may challenge the patience of younger patients, resulting in 10 patients being excluded from the normative range for this reason. Neural network-based noise cancellation is anticipated in the near future, which may mitigate test noise in practice and facilitate testing in younger children. Caution is also needed if the percentage of the meal consumed in a clinical test is low (<50%).^35^ Specifically, while a very low meal intake does not necessarily preclude interpretation, it may lead to false positives for low amplitude and/or low GA-RI values, particularly in the later post-prandial hours.^23^ The impact of a lower kCal meal on BSGM metrics in adolescents is currently being evaluated. Finally, specific reference intervals are also desirable for younger children, who may find the current protocol challenging. Therefore, test duration, meal volume and composition, and app-based symptom logging procedures will all require further consideration.

By providing the first validated normative dataset for this population, this study represents an important diagnostic step forward for investigating chronic gastroduodenal symptoms in adolescents.

## Data Availability

Data sharing and data use are governed by the Aotearoa | New Zealand Health and Disability Ethics Committee. The majority of the data is available in the manuscript; however, all requests for additional data can be made to the corresponding author. Requests will be granted if the proposed use aligns with the ethical approval for the study and relevant ethical approvals have been obtained, including approval from a New Zealand Ethical Committee and the Childrens Hospital of Philadelphia IRB.  

---

**Supplementary Figure 1.**
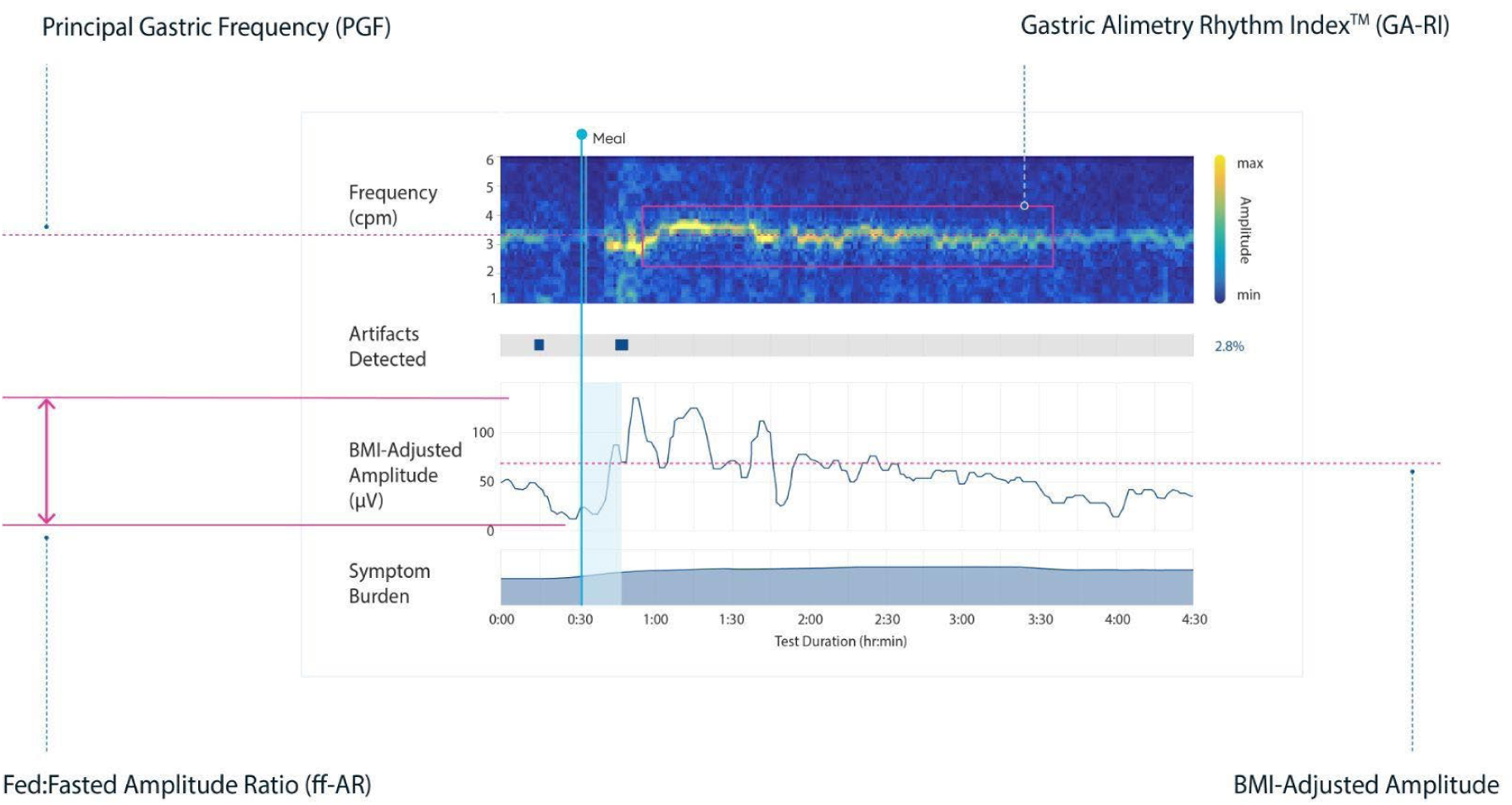
Visualising the Body Surface Gastric Mapping metrics - PGF, BMI-adjusted amplitude, GA-RI and ff-AR - and including artifacts detected and symptom burden using a spectral plot for a single participant.

**Supplementary Figure 2.**
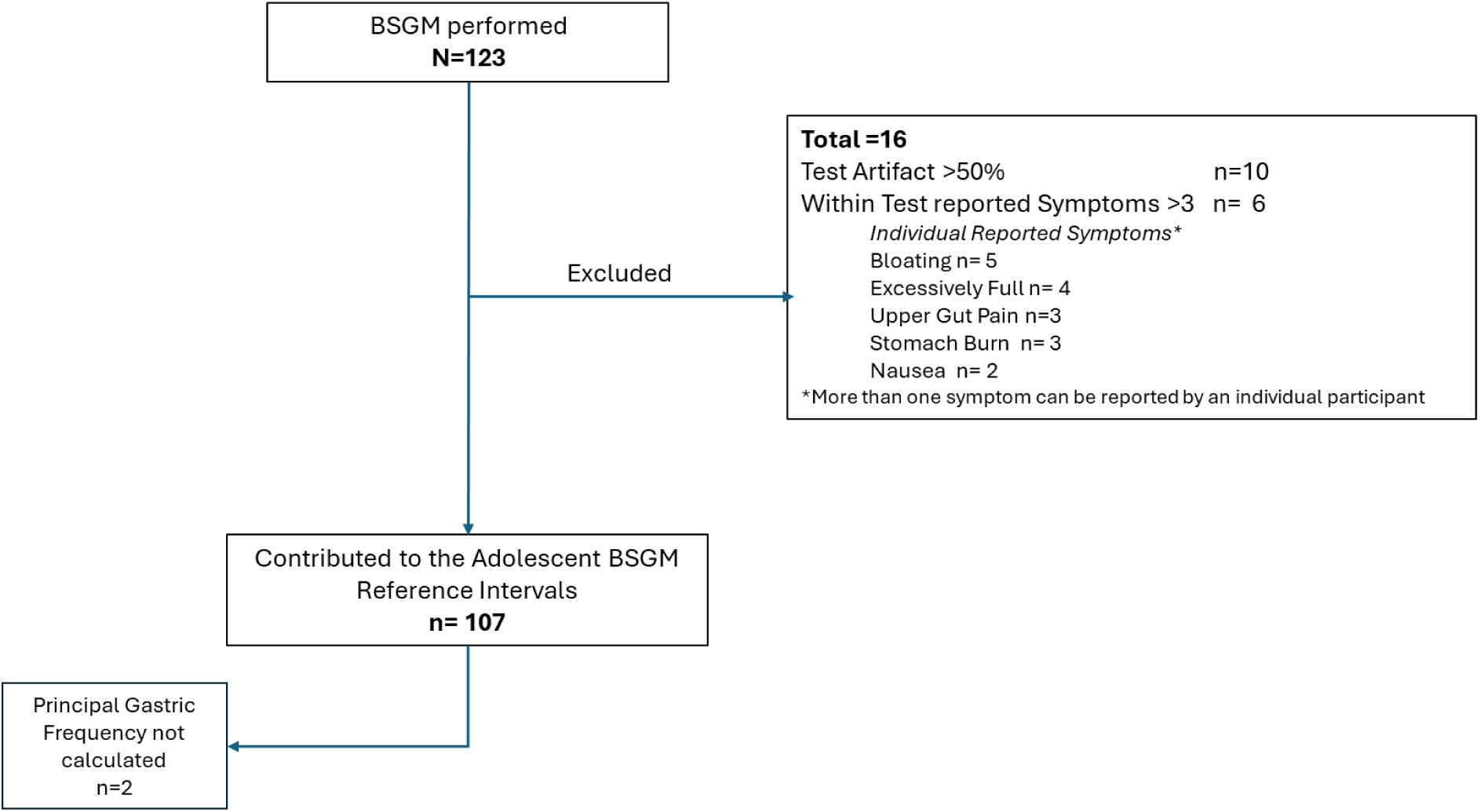
Recruitment flow diagram showing the number of participants recruited, those excluded with reason, and the number of participants contributing to the reference intervals, with exclusions from individual metric reference interval calculations.

**Supplementary Figure 3.**
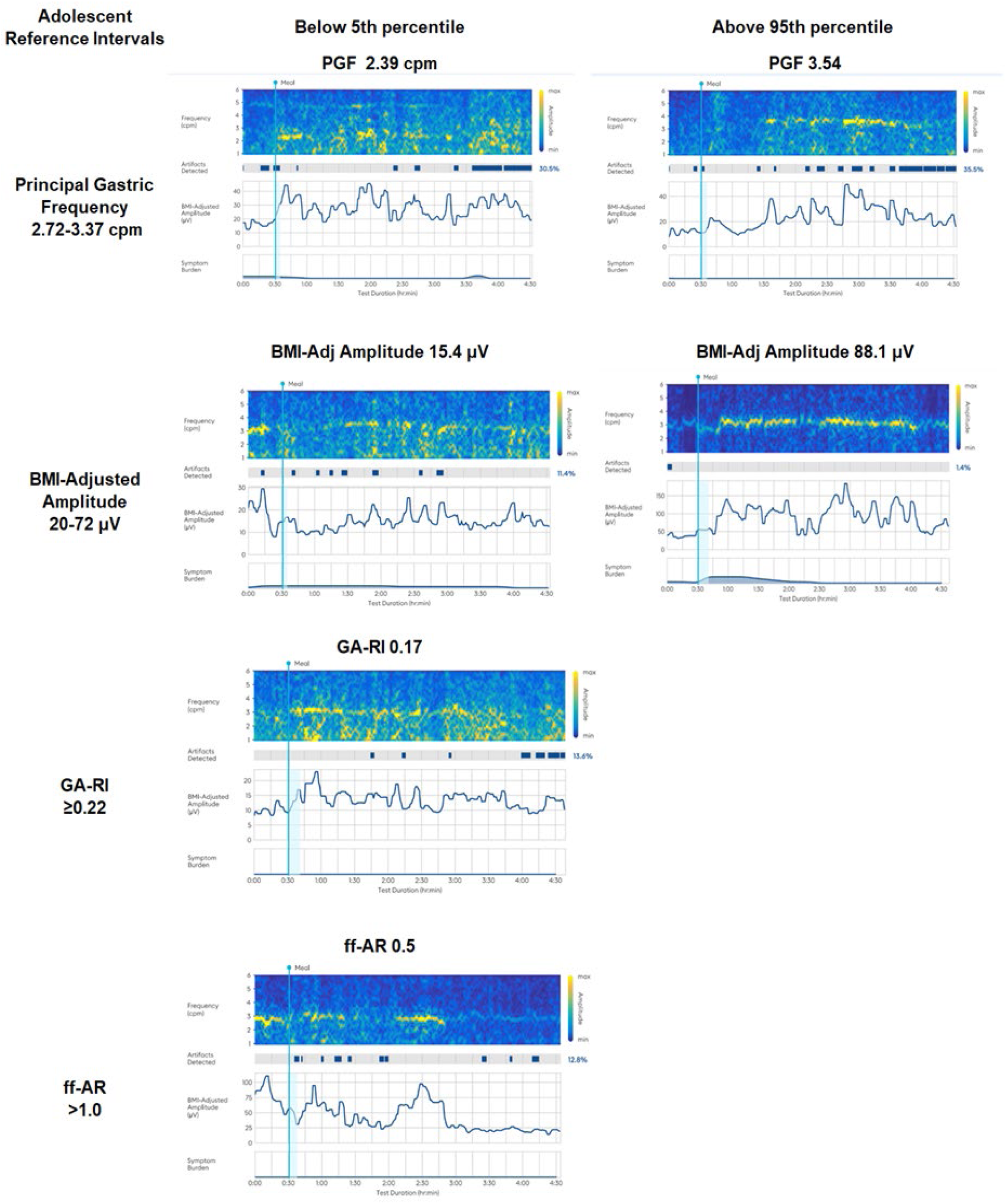
A sample of adolescent healthy control spectrograms that fell outside of the reference intervals for each metric [5th and 95th percentiles for PGF and BMI-adjusted amplitude and 5th percentile for GA-RI and ff-AR]

**Supplementary Table 1.** Univariate and multivariate models of the impact of participant demographic characteristics (ethnicity, sex and age) and anthropometric variables (BMI and height) and BSGM metrics (PGF, BMI-adjusted amplitude, GA-RI and ff-AR). The test statistic for the univariate models is either a standardised regression coefficient (age, BMI), t-statistic (sex) or F-statistic (ethnicity). For the multivariate regression model, sex is referenced to male.

| BSGM Metrics | Participant Characteristics | Univariate |  | Multivariate |  |
| --- | --- | --- | --- | --- | --- |
|  |  | Test Statistic (CI) | adj p value | Coefficient (CI) | p value |
| Principal Gastric Frequency | Ethnicity | 0.355 | 0.88 |  |  |
|  | Sex | 1.355 | 0.18 |  |  |
|  | Age (years) | -0.26<br>(-0.42 – -0.10) | 0.007 | -0.16<br>(-0.34 – 0.02) | 0.154 |
|  | BMI (kg/m<sup>2</sup>) | -0.16<br>(-0.32 – -0.00) | 0.09 |  |  |
|  | Height (cm) | -0.29<br>(-0.44 – -0.13) | 0.003 | -0.21<br>(-0.39 – -0.02) | 0.063 |
| Log BMI-Adjusted Amplitude | Ethnicity | 1.191 | 0.319 |  |  |
|  | Sex | 1.03 | 0.304 |  |  |
|  | Age (years) | 0.02<br>(-0.14 – 0.19) | 0.81 |  |  |
|  | BMI (kg/m<sup>2</sup>) | -0.27<br>(-0.43 – -0.11) | 0.005 |  |  |
|  | Height (cm) | -0.11<br>(-0.28 – 0.05) | 0.24 |  |  |
| Gastric Alimetry Rhythm Index | Ethnicity | 1.743 | 0.132 |  |  |
|  | Sex | 0.958 | 0.34 |  |  |
|  | Age (years) | 0.16<br>(0.00 – 0.32) | 0.093 |  |  |
|  | BMI (kg/m<sup>2</sup>) | -0.21<br>(-0.37 – -0.05) | 0.029 |  |  |
|  | Height (cm) | 0.02<br>(-0.14 – 0.18) | 0.856 |  |  |
| Log Fed-Fasted Amplitude Ratio | Ethnicity | 0.439 | 0.821 |  |  |
|  | Sex | -1.102 | 0.273 |  |  |
|  | Age (years) | -0.20<br>(-0.36 – -0.04) | 0.042 | -0.06<br>(-0.24 – 0.12) | 0.572 |
|  | BMI (kg/m<sup>2</sup>) | -0.27<br>(-0.42 – -0.11) | 0.005 | -0.22<br>(-0.38 – -0.06) | 0.026 |
|  | Height (cm) | -0.23<br>(-0.39 – -0.08) | 0.016 | -0.16<br>(-0.34 – 0.02) | 0.148 |
| CI, confidence interval; adj, adjusted p value; BMI, body mass index, cm, centimetres |  |  |  |  |  |

**Supplementary Table 2.**
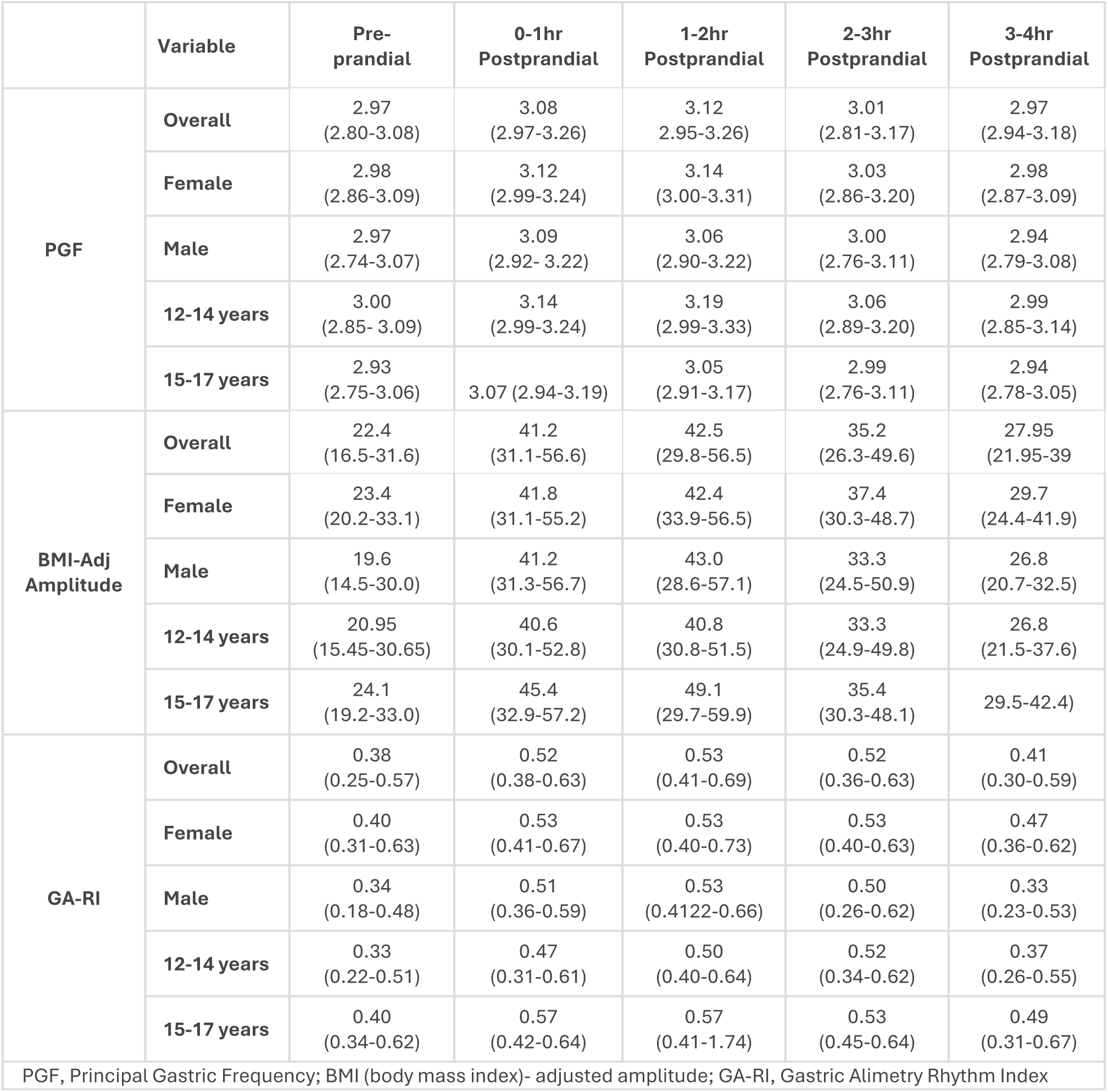
The median (IQR) values for each BSGM metric reported by sex and 12-14 and 15-17 age cohorts, by test time periods

**Supplementary Table 3.** Cross-validation presenting the percentage of adolescent participants in each fold that fell outside the reference intervals compared to the other 4 folds

| Fold Number | Low BMI-Adj Amplitude | High BMI-Adj Amplitude | Low Principal Gastric Frequency | High Principal Gastric Frequency | Low Gastric Alimetry Rhythm Index | Low Fed Fasted Amplitude Ratio | Mean (Std. Dev.) |
| --- | --- | --- | --- | --- | --- | --- | --- |
| 1 | 0 | 9.09 | 18.18 | 9.09 | 4.55 | 4.55 | 7.58 (6.21) |
| 2 | 9.52 | 9.52 | 0 | 0 | 0 | 9.52 | 4.76 (5.22) |
| 3 | 0 | 4.76 | 4.76 | 9.52 | 4.76 | 0 | 3.97 (3.58) |
| 4 | 4.76 | 0 | 0 | 0 | 4.76 | 9.52 | 3.17 (3.89) |
| 5 | 9.52 | 4.76 | 0 | 4.76 | 9.52 | 4.76 | 5.56 (3.58) |
| Mean (Std. Dev.) | 4.76 (4.76) | 5.63 (3.88) | 4.59 (7.87) | 4.68 (4.66) | 4.72 (3.37) | 5.67 (4.00) | 5.01 (1.69) |

